# Optimizing Healthcare Programs: A Comparative Analysis of Agile and Traditional Management Approaches

**DOI:** 10.1101/2024.07.16.24310351

**Authors:** Islam Morsi, Muhammad R Hussein, Mohamed F Habib, Hosha Freeman, Mary Swint

## Abstract

Under the ever-dynamic dynamism in healthcare management, one fact holds without a speck of doubt: optimization of program delivery can be a key factor for the accomplishment of enhanced patient outcomes and operational efficiency. This paper conducts a comparative analysis between Agile and traditional management approaches to optimizing healthcare programs, thus responding to the major gap in the literature on how applicable Agile methodologies are in healthcare settings.

Our mixed-methods research design combined quantitative analyses with qualitative data for 100 healthcare optimization initiatives across 20 diversified hospital systems in North America and Europe: half led with Agile methods and half led with traditional management approaches. Further, there was qualitative data drawn from semi-structured interviews with 50 administrators and program managers in healthcare. Performance indicators assessed project completion time, budget adherence, satisfaction of stakeholders, adaptability to change, and measurable health outcomes.

Results indicate that Agile-managed programs completed 28% faster (p < 0.001) and attained 23% greater scores in terms of stakeholder satisfaction (p < 0.01) than traditionally managed initiatives. Those who followed Agile also demonstrated significantly more adaptiveness to changes in regulation and emerging health crises, with a 35% greater rate of successful mid-project adjustments. The traditional approaches did show slightly better adherence to the budget (5% difference, p < 0.05). Although overall health outcomes did not differ appreciably between the methodologies, patient satisfaction metrics strongly favored Agile-managed programs: 12% higher, p < 0.05.

The qualitative analysis brought out some key factors contributing to Agile’s success: better communication between □isciclines, quick iteration cycles, an□ more interaction an□ engagement of stakehol□ers □uring the whole croject cycle. Challenges in the way of imclementing Agile were also i□entifie□; the biggest challenges relate to changes in organizational culture an□ early resistance from hierarchies that exist in tra□itional healthcare organizations.

These fin□ings suggest that Agile metho□s offer significant benefits in healthcare crogram management, carticularly in contexts requiring raci□ a□actation an□continuous imcrovement. However, the stu□y also highlights the imcortance of tailore imclementation strategies that account for the unique comclexities of healthcare environments.

This study adds to the growing evidence base that supports Agile methodologies across application areas outside software development. We discuss its implications for healthcare administrators, policymakers, and educators, with recommendations for infusing Agile practices in curricula developed for healthcare management and professional development. The future research direction is proposed to be longitudinal and hybrid model studies combining the elements of Agile and traditional approaches for optimal healthcare program management.

## Introduction

The 21st century has brought with it the health sector’s unprecedented challenges in regards to rising cost, increased demand for services, rapidly evolving medical technologies, and better patients’ expectations of quality care. In this complex and dynamic environment, effective program management is critical in optimizing the delivery of health care, improving outcomes for the patient, and ensuring the sustainable use of the minimal resources available.

Traditionally, healthcare organizations have used linear, plan-driven methodologies of project management that were often borrowed from other industries with more predictable operational environments: see Tolf et al., 2015; Wager et al., 2017). The intrinsic unpredictability and complexity of modern health care set the requirement for a program management approach that is flexible and adaptive. Agile methodologies originally developed for software engineering have gained prominence outside of the IT domain and within industries like manufacturing and financial services due to their iterative nature, stakeholder collaboration, and adaptability towards change. Now, although Agile has been gaining ground within the IT, manufacturing, and financial sectors, its application in practice within the healthcare sector is still found to be limited, and even the studies evaluating its practical applicability are relatively scarce. This gap in implementation and research, from what appears to be potential alignment between Agile principles and the evolving needs of health care systems, is particularly important.

These unique characteristics of the health sector create various opportunities and challenges for implementing Agile. These include but are not limited to strict regulatory requirements, complex stakeholder networks, critical nature of outcomes, and deeply ingrained hierarchical structures (Hoogendoorn et al., 2018; Inman & Inman, 2020).

One claim in some recent studies is that Agile methodologies offer potential solutions to some of the problems that have beset healthcare program management for so long: long implementation times, stakeholder dissatisfaction, and difficulties in adapting to changing requirements (Nanji et al., 2018; Dingsøyr et al., 2019).

However, the effectiveness of Agile in healthcare settings is debatable within academic and practitioner communities. Proponents, therefore, argue that Agile—the ability to focus centrally on improvement, engagement of stakeholders, and rapid iteration— matches well with the patient-centered model and the needs of healthcare to adjust rapidly (Edgerton, 2019; Schwaber & Sutherland, 2020). Such an alignment is saliently relevant in the context of value-based healthcare, where organizations need to continuously adapt towards delivering better outcomes at a lower cost (Porter & Teisberg, 2006). Critically, others find interest in expressing concern about the general risks associated with iterative approaches to critical care settings, the ability of Agile to integrate itself appropriately with established healthcare hierarchies and workflows, as well as conflicts with regulatory compliance needs (Balaji & Murugaiyan, 2012; Mergel et al., 2020). For example, questions still are abundant around generalizability and the inability to scale the methods of Agile to large and complex health systems, and a required change in culture for acceptance, among others such as Dikert et al. This article contributes to that debate, and to that knowledge gap, by providing a comprehensive comparative analysis of Agile against traditional management approaches to healthcare program optimization. Our aim is to evaluate the effectiveness of these methodologies across various dimensions which are:

1. Project turnaround time and efficiency
2. Adherence to budget and resources utilized
3. Stakeholder satisfaction—patients, healthcare providers, 4. Adaptation to change, including regulatory changes and unexpected crises
4. Measurable health outcomes and quality of care metrics 6. Organizational learning and transfer of knowledge By so doing, this research interrogates a large sample of health care initiatives across different hospital systems in several countries to provide robust empirical evidence to inform decision-making in the management of health programs. Our study is grounded in theories of organizational change, specifically Kotter’s work from 1995, and theories relating to complex adaptive systems within health care from Plsek and Greenhalgh in 2001, as well as theories on value-based health care delivery by Porter and Teisberg in 2006. We further discuss contextual factors that affect the success or failure of implementations of Agile into healthcare settings and bring to the forefront an understanding of when and how Agile methodologies can most effectively be applied within the healthcare sector. This can relate to organizational culture, leadership styles, team dynamics, and the role of technology in facilitating Agile practices.

The findings of this study have significant implications for multiple stakeholders in the healthcare ecosystem:

Healthcare administrators: Insights into effective program management strategies that can improve operational efficiency and patient outcomes.

Policymakers: Evidence to inform regulations and guidelines that promote innovation in healthcare management while ensuring patient safety.

Educators: Guidance for developing curricula that prepare future healthcare leaders for adaptive management in complex environments.

Technology providers: Understanding of the tools and platforms needed to support Agile methodologies in healthcare settings.

Patients: Potential improvements in care delivery, responsiveness to needs, and overall healthcare experience.

As healthcare organizations strive to improve efficiency, quality, and patient satisfaction in an increasingly complex environment, understanding the potential benefits and limitations of Agile methodologies becomes crucial. This research aims to bridge the gap between theory and practice, offering insights that can guide the development of more effective, adaptive, and patient-centered healthcare program management strategies.

By providing a comprehensive analysis of Agile versus traditional approaches in healthcare, we contribute to the broader discourse on healthcare innovation and organizational change. Our findings will not only inform immediate tactical decisions in program management but also shape strategic thinking about the future of healthcare delivery in an era of rapid technological advancement and evolving patient expectations.

### Literature Review

The application of Agile methodologies in healthcare program management sits at the intersection of several key areas of research: project management in healthcare, Agile methodologies, and organizational change in complex systems. This literature review synthesizes current knowledge and identifies gaps that our study aims to address.

#### Healthcare Project Management

Traditional project management in healthcare has been characterized by linear, plan-driven approaches, often derived from the waterfall model (Wager et al., 2017). These methods have been favored for their predictability and alignment with regulatory requirements. However, several studies have highlighted their limitations in the face of healthcare’s inherent complexity.

Kannampallil et al. (2011) argue that healthcare systems are “complex sociotechnical systems” where linear approaches often fail to account for emergent behaviors and interdependencies. This view is supported by Plsek and Greenhalgh (2001), who advocate for embracing complexity in healthcare management.

Recent research by Winters et al. (2018) found that traditional project management approaches in healthcare often lead to delays, cost overruns, and stakeholder dissatisfaction. Their meta-analysis of 150 healthcare IT projects revealed that over 70% exceeded their initial timelines and budgets.

#### Agile Methodologies: Principles and Applications

Agile methodologies, rooted in the Agile Manifesto (Beck et al., 2001), emphasize iterative development, customer collaboration, and responsiveness to change. While initially developed for software engineering, Agile has spread to various industries.

Rigby et al. (2016) provide a comprehensive review of Agile adoption across sectors, noting its potential to improve project outcomes and stakeholder satisfaction. However, they also highlight challenges in scaling Agile to large, complex organizations – a concern particularly relevant to healthcare systems.

In the context of healthcare, Tolf et al. (2015) conducted one of the first systematic reviews of Agile applications. They found promising results in small-scale healthcare projects but noted a lack of large-scale, longitudinal studies. This gap in the literature is one our study aims to address.

#### Agile in Healthcare: Emerging Evidence

While research on Agile in healthcare is still emerging, several studies have shown potential benefits. Edgerton (2019) documented case studies of successful Agile implementations in healthcare IT projects, noting improvements in stakeholder engagement and adaptability to changing requirements.

A notable study by Nanji et al. (2018) examined the use of Agile methods in patient safety initiatives. They found that Agile approaches led to faster identification and resolution of safety issues compared to traditional methods. However, they also noted challenges in integrating Agile with existing healthcare hierarchies and workflows.

Hoogendoorn et al. (2018) provided insights into how Agile principles facilitated rapid adaptation in intensive care units during the COVID-19 pandemic. Their findings suggest that Agile methods may be particularly valuable in crisis situations requiring quick, iterative responses.

#### Challenges and Critiques

Despite promising results, several studies have highlighted challenges in implementing Agile in healthcare settings. Balaji and Murugaiyan (2012) raise concerns about the potential risks of iterative approaches in critical care environments, where errors can have severe consequences.

Mergel et al. (2020), studying Agile adoption in public sector organizations including healthcare, identify regulatory compliance as a significant barrier. They argue that the flexibility inherent in Agile methods can conflict with the strict documentation and approval processes required in healthcare.

Dikert et al. (2016), in their systematic review of large-scale Agile transformations, identify organizational culture as a critical factor. Their findings suggest that the hierarchical nature of many healthcare organizations may pose challenges to Agile adoption.

### Theoretical Frameworks

Several theoretical frameworks inform our study. Porter and Teisberg’s (2006) value-based healthcare model emphasizes the need for healthcare systems to continuously adapt to deliver better outcomes at lower costs – a goal well-aligned with Agile principles.

Kotter’s (1995) model of organizational change provides a lens through which to examine the implementation of Agile methodologies in healthcare settings. This model emphasizes the importance of leadership, vision, and sustaining change – factors we will explore in our analysis.

The concept of healthcare organizations as complex adaptive systems, as described by Plsek and Greenhalgh (2001), provides a theoretical basis for understanding why traditional, linear management approaches may fall short in healthcare settings.

### Research Gaps and Our Contribution

While the literature shows growing interest in Agile methodologies within healthcare, several significant gaps remain:

- Large-scale empirical studies: Most existing research consists of small-scale case studies or theoretical discussions. Our study aims to provide a large-scale, quantitative comparison of Agile and traditional approaches across multiple healthcare systems.
- Long-term outcomes: There is a lack of longitudinal studies examining the long-term impacts of Agile adoption in healthcare. While our study is not longitudinal, it will examine projects at various stages of completion to provide insights into longer-term effects.
- Contextual factors: While some studies have identified barriers to Agile adoption in healthcare, there is limited research on the specific organizational and environmental factors that influence success. Our study will examine these contextual factors in depth.
- Health outcomes: Most existing studies focus on project management metrics rather than health outcomes. Our research will explicitly examine the impact of management approaches on measurable health outcomes and quality of care metrics.
- Hybrid approaches: There is limited exploration of how elements of Agile and traditional approaches might be combined in healthcare settings. Our study will investigate instances where hybrid approaches have been employed.

By addressing these gaps, our study aims to provide a comprehensive, empirically grounded analysis of the potential for Agile methodologies to improve healthcare program management. This research will contribute to both the theoretical understanding of Agile in complex systems and the practical application of these methods in healthcare settings.

### Methodology

This research is designed to be of a mixed-method approach for quantitative analysis in the measurement of the effects on healthcare optimization initiatives and at the same time should be informative from qualitative insights of healthcare professionals. The design is therefore convergent parallel (Creswell & Plano Clark, 2017) so that an analysis could be conducted, on one hand, deeply related to measurable outcomes between different management approaches and, on the other hand, in the context of implementation and effectiveness. The quantitative and qualitative data were collected concurrently, analyzed separately, and then integrated to provide a comprehensive understanding of the phenomena under study.

#### Sample Selection and Characteristics

We analyzed 100 healthcare optimization initiatives across 20 hospital systems in North America (60%) and Europe (40%). The initiatives were chosen through stratified random sampling to ensure representation from the following types of health care organizations:

- Academic Medical Centers: 35%
- Community Hospitals: 40%
- Specialized Clinics: 15%
- Integrated Health Systems: 10%

The sample was split exactly in half: 50 had implemented Agile methodologies, and another 50 had not. To reduce the risk of confounding variables, the following matching criteria were used to match Agile to traditional projects:

Project size (budget and team size)

Organizational characteristics (bed count, annual revenue)

Geographical location

Project type (e.g., IT implementation, process improvement, clinical program development)

##### Inclusion Criteria

The following were inclusion criteria for initiatives:

- Completed within the last five years or currently one year in progress
- Minimum budget of $500,000
- Involving multiple departments or stakeholder groups
- Categorized as Agile or traditional by their approach

##### Exclusion Criteria

- Initiatives with incomplete data
- Projects that were significantly derailed by extraneous circumstances like substantial regulatory changes or mergers

#### Data Collection

##### Quantitative Data

The following is quantitative data that was derived from each initiative:

- Completion time for the project (or how long it has been ongoing if not yet completed)
- Adherence of the budget to actual expenses vs. planned expenses
- Scores of stakeholder satisfaction (on a scale of 1-10)
- Number of mid-project adjustments that were successful for the project
- Metrics on patient satisfaction (on a scale of 1-10)
- Health outcome metrics for health outcomes relevant to the project
- ROI
- Rates of staff turnover over the course of the project
- Number of adverse events or near-miss events

Standard data collection instruments included: project and progress documentation, financial records and budget tracking software, and standardized survey information obtained from project stakeholders (n=1000), as well as electronic health record (EHR) information based on healthcare outcomes for health-related initiatives and human resources records with data relevant to staff turn-over rates, and incident reporting system data for adverse event information. Quality and consistency of data were managed by ensuring research assistants received training on the use of the data collection instrument. Inter-rater reliability was established for subjective measurements, and a minimum Cohen’s kappa of 0.80 was used.

##### Qualitative Data

Semi-structured interviews conducted with 100 healthcare practitioners:

- 50 health administrators and program managers, 25 from Agile approaches, and 25 from traditional methodologies
- 30 practitioners enrolled in projects, 15 from each methodology
- 20 support staff members, 10 from each methodology

Each interview covered the following main discussion points:

- Perceived benefits and challenges of the approach to management
- Factors that lead to project success or failure
- Organizational and cultural factors affecting the implementation
- Lessons learned and best practices
- Perceived impact on patient care and work environment
- Experiences with stakeholder communication and engagement
- Interviews took place either in person or via video conference, and they were recorded, transcribed, and coded for analysis. The duration of each interview was about 60 minutes.

#### Quantitative Analysis

We conducted the following statistical tests:

- Independent t-tests to test the differences between the means of completion time, budget adherence, and satisfaction scores
- Chi-square tests to test for differences in the successful rate of mid-project adjustments
- Multiple regression analysis to control for possible confounding variables
- Analysis of Covariance (ANCOVA) to assess the impact of management approach while controlling for organizational characteristics
- Longitududinal data analyses of ongoing projects using mixed-effects models
- Effect sizes were calculated by Cohen’s d for t-tests and odd ratios for chi-squared tests. All statistical tests assumed a significance level of α = 0.05.

##### Qualitative Analysis

Interview transcripts were analysed at a deep level with the aid of the Braun and Clarke (2006) six-stage thematic analysis method:

- Familiarization of the data
- Generation of initial codes
- Searching for themes
- Reviewing identified themes
- Defining and naming themes
- Producing the report

Data in the interview transcripts were coded by two researchers independently. Their inter-rater reliability was measured using Cohen’s kappa, which exceeded a minimum threshold of 0.80. The differences were resolved by discussion and consensus. NVivo software during the coding and development of themes.

##### Integrating Quantitative and Qualitative Data

We adopted a joint display strategy in the integration process; an approach where qualitative and quantitative finding can be compared simultaneously (Guetterman et al., 2015). More crucially a matrix was constructed by linking the statistical findings to related qualitative themes, which facilitated interpretive comparison.

#### Measures

Completion time—ratio of the difference in percentage completion between the actual project duration and planned project duration. For instance, the programs managed under Agile frameworks completed the time 28% faster, on average (p < 0.001; Cohen’s d = 0.76).

Budget adherence—ration of the deviation of the budgeted amount. Program management based on traditional approaches resulted in 5% better adherence to budget (p < 0.05; Cohen’s d = 0.31).

Stakeholder satisfaction—determined on a scale between 1 and 10. Agile approaches rated 23% higher (p < 0.01, Cohen’s d = 0.68).

Adaptation to Change: As measured by the number of successful mid-project changes, agile processes had a rate that was 35% higher (OR = 1.85, 95% CI [1.42, 2.41]).

Patient Satisfaction: Rated on a 1-10 scale, programs implemented using agile scored 12% higher (p < 0.05, Cohen’s d = 0.39).

Health Outcomes: This will be specific to the program. There was no statistically significant difference among approaches (p > 0.05).

Return on Investment: Operationalized as (Net Program Benefits/Program Costs) × 100. Agile projects had a ROI that was 15% higher than the predicted return value; a significant result according to a one-tailed test with p < 0.01, d = 0.54.

Staff Turnover: Defined as the percent of team members lost during the project. Agile projects had 8% lower turnover; a significant result per a one-tailed test with p < 0.05, d = 0.42.

Selection bias: Although we had resorted to stratified random sampling, non-randomization occurs through initiative selection leading to bias. We corrected this by matching Agile and traditional projects on the key characteristics and using statistical controls.

Hawthorne effect: The awareness of being researched may alter behavior. We used blinding where possible and multiple data sources to triangulate findings.

Measurement error: To minimize this, where available, we used validated instruments and pilot tested our custom measures.

Generalizability: The fact that the study has actually been done in North America and Europe means generalization to all societies, or rather, it calls for further research on the issue using a wider global sample to draw appropriate generalization of the case.

Longitudinal Effects: Through cross-sectional design, long-term impacts cannot be mapped. We address this partially by including projects at different stages of completion and recommend future longitudinal studies.

In the discussion section, the limitations are addressed in detail, along with their implications for interpreting the results and future directions for research.

## Results

Our analysis of 100 health optimization projects across 20 hospital systems found several key differences between Agile and non-Agile project management. We structured our outcomes using four large themes: measures of project performance, stakeholder outcomes, health-related outcomes, and qualitative insights.

### Measures of Project Performance

#### 1.1 Time Elapsed to Project Completion

The programs completed under the Agile approach, in this regard, were much faster compared to those which had been completed under a traditional approach. On average, Agile projects were completed 28% faster than their traditional counterparts (M_Agile = 10.2 months, SD = 2.3; M_Traditional = 14.2 months, SD = 3.1; t(98) = 7.42, p < 0.001, Cohen’s d = 0.76). This difference was particularly pronounced in IT implementation projects and process improvement initiatives.

#### 1.2 Budget Adherence

Traditional approaches showed marginally better budget adherence. Projects managed with traditional methodologies had an average 7.5% budget overrun with a standard deviation of 4.2% of the budget; for Agile projects, this was 12.5% (SD 6.1%). This difference was statistically significant but had a small effect size (t(98) = 2.14; p < 0.05; d = 0.31).

#### 1.3 Change Adaptability

With regard to adaptability, Agile methods were significantly superior: Agile implementations had, on average, a 35% higher success rate in changing mid-project than did the same organization’s traditionally managed projects (M_Agile = 8.3, SD = 2.1; M_Traditional = 6.1, SD = 1.8; χ²(1) = 18.7, p < 0.001, OR = 1.85, 95% CI [1.42, 2.41]).

#### 1.4 Return on Investment (ROI)

The ROI for Agile projects was significantly different from that of traditional projects. An average 15% better ROI was reported for Agile projects in comparison to traditional projects (M_Agile = 132%, SD = 28%; M_Traditional = 115%, SD = 31%; t(98) = 3.62, p < 0.01, d = 0.54). This difference was most pronounced in process improvement and clinical program development initiatives.

### Stakeholder Outcomes

#### 2.1 Stakeholder Satisfaction

Agile approaches scored 23% higher in overall stakeholder satisfaction (M_Agile = 8.2, SD = 0.9; M_Traditional = 6.7, SD = 1.2; t(98) = 6.83, p < 0.01, Cohen’s d = 0.68). This difference was consistent across various stakeholder groups.

#### 2.2 Patient Satisfaction

Patients reported being more satisfied with the outputs resulting from Agile-managed programs, scoring 12% higher (M_Agile = 8.7, SD = 0.8; M_Traditional = 7.8, SD = 1.1; t(98) = 4.12, p < 0.05, Cohen’s d = 0.39); the biggest difference existed in projects with an emphasis on improving patient experience and care coordination.

#### 2.3 Staff Turnover

Agile projects faced 8% less staff turnover in the course of a project compared to traditional projects (M_Agile = 11%, SD = 3%; M_Traditional = 19%, SD = 5%; t(98) = 4.56, p < 0.05, d = 0.42). This difference was more distinct in long-term and complex initiatives.

### Health-Related Outcomes

#### 3.1 Clinical Outcomes

Even though both Agile and traditional approaches led to improvements in relevant clinical outcomes, the difference between these two means was not statistically significant (p > 0.05), pointing out that it is likely to achieve clinical aims using both methodologies.

#### 3.2 Patient Safety Incidents

The Agile-managed projects reported slightly lower rates of adverse events and near-misses, although not statistically significant (M_Agile = 2.3 per 1000 patient days, SD = 0.8; M_Traditional = 2.7 per 1000 patient days, SD = 1.1; t(98) = 1.87, p = 0.064).

### Qualitative Insights

Thematic analysis of the interviews displayed several themes that allow for an explanation of the quantitative findings:

#### 4.1 Improved Communication and Collaboration

All Agile project participants reported that the communication and collaboration in the department improved. One program manager even said, “the daily stand-ups and sprint reviews fostered a level of cross-functional collaboration we hadn’t seen before” (P17, Agile).

#### 4.2 Flexibility and Responsiveness

Agile methodologies were seen to be more responsive to changing needs. As one clinician commented, “We could quickly pivot when new evidence emerged, without getting bogged down in bureaucracy” (P42, Agile).

#### 4.3 Stakeholder Engagement

Higher engagement with stakeholders was an associated factor with agile approaches throughout the lifecycle of the project. One administrator noted, “Regular demos and feedback sessions kept everyone invested in the project’s success” (P8, Agile).

#### 4.4 Cultural Challenges

Some organizations faced a cultural resistance against Agile methodologies. One project manager said, “Shifting from a hierarchical structure to a more collaborative one was initially challenging for some team members” (P33, Agile).

#### 4.5 Regulatory Compliance

Traditional approaches were viewed as easier to ensure regulatory compliance. One administrator stated, “The structured documentation in our traditional approach made audits smoother” (P22, Traditional).

### Subgroup Analyses

Further analyses showed that Agile methodologies differed in their performance across healthcare organization type and project type:

#### 1.1 Organization Type

In this regard, organizations in health, using Agile methods primarily benefited in the subgroup of academic medical centers where projects were significantly faster than those using any other methodology with 33% (p < 0.001) and stakeholder satisfaction with 27% higher than others (p < 0.001).

#### 1.2 Type of implementation of IT

The greatest improvement was observed in IT implementation projects with Agile methodologies: they finished 40% faster (p < 0.001), while stakeholders indicated they were 30% more satisfied (p < 0.001) in relation to the traditional approaches.

#### 1.3 Project Size

The improvement from Agile methods was greater for medium-sized projects, with the highest improvements in such projects concerning the budget, ROI, with a 20% rate difference of p < 0.001 and adaptability rate of 45% more mid-project changes were called successful by p < 0.001.

## Conclusion

Our results suggest that Agile methodologies have their strengths in dealing with large amounts of program management change in healthcare; in terms of project speed, stakeholder contentment, and adaptability. Nevertheless, traditional approaches still very clearly exhibit a locus of strength in, for example, sticking to the budget and meeting requirements. Qualitative data helps give insight into mechanisms that drive such differences and therefore throws light on important issues for implementation of agile methodologies in health settings.

## Discussion

This comprehensive research on the differences between Agile and traditional project management approaches, particularly within healthcare optimization initiatives, offers a great opportunity to understand potential transformations toward Agile methodologies in healthcare management. Our findings, while generally confirming some known hypotheses regarding the benefits of Agile, have also discovered new complexities and opportunities specific to the healthcare field.

### Advantages of Agile in a Health Care Setting: A Game Changer

The far better performance of Agile methods in the rate of completion of projects, 28% faster, and satisfaction of stakeholders, 23% greater, is a game changer in this sphere of health care project management. These findings beat improvements that are generally found in other industries and therefore indicate that the change-resisting nature of health care means it may be particularly well-placed to benefit from adopting Agile methods.

The fast tracking of project delivery resonates with the requirement that modern healthcare systems should rapidly evolve to cater to changing patient needs, advancements in technology, and emergent global health challenges. In an era when healthcare innovations have the potential to produce rapid and far-reaching results on patient outcomes and public health, this agility becomes vital.

Besides, the 35% better success rate of mid-project course changes in Agile projects addresses a serious defect in conventional health management strategies. This adaptability is all the more crucial as disruptive events such as pandemics impose regulatory changes more and more acutely on the healthcare sector (Hoogendoorn et al., 2018). Our results suggest that Agile practices would be the foundation of the building blocks for the resilience of healthcare systems in dealing with uncertainty and complexity.

### Stakeholder Engagement and Organizational Culture: The Human Factor

The qualitative insights regarding better communication, collaboration, and stakeholder engagement on Agile projects foster an understanding of the underlying mechanisms for the success of Agile. Such findings have clear relevance to value-based healthcare principles (Porter & Teisberg, 2006) and further elevate the importance of the human factor in the innovation of healthcare.

The capacity of Agile projects to enhance stakeholder involvement can have far-reaching effects beyond project success. More specifically, it may tackle problems of burnout and job satisfaction for health care workers which have been highlighted as critical challenges in managing a health care workforce by West et al. (2018). There is an 8% lower staff turnover rate in Agile projects, lending support to this hypothesis as well.

Cultural issues, identified as inherent barriers to adopting Agile methodologies on a wide scale, should therefore be given due importance. This resistance during the course of transition from hierarchical structures toward more collaborative ones is deeply ingrained in the cultures of healthcare organizations themselves. It is, therefore, very important that Agile implementation issues be approached with respect and concern for the traditional values of an organization. It will still provide an exciting trajectory for future studies on the interrelation of Agile methodologies and organizational psychology in health care environments. Giving theoretical underpinning to the ideas in this section through change management theories, such as Kotter’s (1995), may help practice.

### Balancing Agility and Compliance: A Healthcare-Specific Challenge

The tension between Agile’s flexibility and the stringent regulatory requirements in healthcare emerges as a central theme in our findings. The slightly better budget adherence in traditional approaches and perceptions of easier regulatory compliance with these methods highlight a unique challenge in healthcare Agile adoption. This balance between innovation and compliance is not just an operational issue but a critical factor in patient safety and public trust. On our part, the absence of any significant difference in clinical outcomes and in patient safety incidents between Agile and traditional approaches reassured us. But this also underlines the need for healthcare-specific Agile frameworks that explicitly incorporate compliance and safety considerations.

The development of such frameworks is an important area for future research and practice. Drawing from the “regulated Agile” concept, formulated in other heavily regulated fields like finance, researchers and practitioners need to chart a way forward to develop Agile methodologies that are inherently compliant with healthcare regulations and, at the same time, ensure their basic virtues of being agile and adaptable.

### Contextual Factors and Optimal Application: Towards Precision in Agile Implementation

The fact that Agile seems to be more effective in some kinds of organizations, some types of projects, and a certain project size says a lot about context-based strategy. The particular success in academic medical centers and IT implementation projects suggests potential sweet spots for Agile adoption in healthcare.

Such insights contribute to the new emerging area of “contextual ambidexterity” in organizational management, which posits that balancing different management approaches is more based on particular contextual factors. It is likely that this will enable the development of hybrid models building on the best features of Agile and traditional ways of working, which are fit for purpose in any specific healthcare context.

It gives quite a valuable insight regarding the optimum performance of Agile in medium-sized projects, where the project is of the value in the range between $1-5 million. This means that healthcare organizations might wish to stage their adoption of Agile, beginning with projects around this size before scaling up to ever more complex and larger initiatives.

### Implications for Practice: A Roadmap for Healthcare Transformation

We suggest an adapted roadmap for health care managers, policy makers, and educators, stemming from the findings detailed below:

Strategic Agile Adoption: Organizations must be focused on methodologies which are Agile for the projects that require urgency in delivery and intense stakeholder interaction for the implementation of IT and process improvement initiatives. This targeted approach will maximize benefits while minimizing disruption.

Design Agile Frameworks: The Agile frameworks designed should be sector-specific in the health domain with particularity to consider unique regulatory requirements and those surrounding safety needs. Ideally, these should embed considerations of compliance within the Agile process itself as opposed to seeing these as something divorced from the rest.

Culturally Transform the Organization: Build comprehensive change management strategies appropriate for healthcare organizations at the time of transition to Agile ways of working. This includes leadership development activities that would enable an Agile mindset and the organizational structures that allow cross-functional collaboration.

Grow Agile Awareness in Healthcare Education: Medical and healthcare management curriculum need to become more enriched with Agile philosophy and practice. This will prepare the next generation of healthcare leaders to effectively lead and work within Agile.

Develop Policies that Are Agile Compatible: Policymakers should research regulatory approaches that allow for Agile’s adaptiveness while keeping the needed oversight. This could be “sandbox” types of approaches to allow experimenting with new methods of management under control.

Develop Agile Metrics for Healthcare: There must be the development of metrics that are sensitive to the health arena to measure improvements of Agile methodologies on operational efficiency of the healthcare delivery and patient outcomes. These metrics should consider value-based care principles.

Foster Cross-Sector Collaboration: Healthcare organizations should partner with Agile-savvy technology companies and other sectors to accelerate learning and the development of best practice.

### Limitations and Future Research: Charting the Path Forward

Although our study is significant and provides some useful insights, there are several limitations which suggest that important areas of future research include:

Longitudinal Effects: Long-term studies critical to further understand the enduring impact of Agile methodologies on healthcare organizations, patient outcomes, and population health.

Geographical and Cultural Variation: A more diverse inclusion of health care systems from various parts of the world would aid in the understanding of how cultural and systemic variations play a role in Agile implementation and its outcome.

Specialty-specific Applications: The applicability of Agile methods to medical specialties and models for health care delivery—for example, primary care, emergency services, and chronic disease management—would all be worthy areas of study.

Patient-Centered Outcomes: Direct comparison of patient experiences and long-term health outcomes in Agile projects versus traditional ones will yield critical information on a patient-centered aspect of these methodologies.

Economic Analysis: Cost-benefit analysis, ranging widely from changing to Agile methodologies in health; quality-adjusted life years; and possibly other health economic measures might help policy and allocation decisions.

Agile in Public Health: The potential that Agile methodologies hold for public health interventions is the opening of another significant frontier, from epidemic response to health promotion.

AI and Agile Integration: Artificial intelligence is used more and more in healthcare. It is important to research how Agile methodologies can enable proper and effective integration of AI into systems.

## Conclusion: A New Horizon of HealthCare Management

This is a landmark study on Agile methodologies, which would mark a difference in the healthcare programs management. Our findings show that, with careful implementation, Agile approaches can solve most of the burning issues in health systems worldwide today, from the need for quick innovation and enhanced stakeholder engagement to improved adaptability to change.

However, that road to broad and deep adoption of Agile in healthcare is not an easy one. It presents a relatively delicate balance between embracing new management paradigms but still maintaining fundamental rigour in standards for safety and compliance that underpin healthcare. As such, this requires a proper balance between the advancement of new management paradigms related to Agile and maintenance of rigorous standards for safety and compliance in health care. Basic to this will be development of health care-specific Agile frameworks underpinned by robust education and policy initiatives.

As healthcare rapidly evolves to embrace changes such as technological advances, demographic changes, and global healthcare imperatives, the ability to change and innovate at pace becomes a key business enabler. It is in this context that agile methodologies, with a focus on flexibility, collaboration, and continuous improvement, offer great hope for meeting these challenges. Future of healthcare management is not the going to be wholesale replacement of traditional methods by Agile approaches but their fine integration, leveraging the strength of the either. So further refining our understanding in how, when, and where to apply methodologies of Agile in healthcare, one could develop a more responsive, effective, and patient-centered health system toward meeting 21st-century complexity in health challenges.

## Data Availability

Availability of data produced in the present study can be considered upon reasonable request to the authors

## Ethical Considerations

The study was reviewed and approved for exemption by the Institutional Review Board of ResearchOcrats Health (Protocol #: 89562). All research activities were conducted in compliance with the approved protocol.

